# Through The Back Door: Expiratory Accumulation Of SARS-Cov-2 In The Olfactory Mucosa As Mechanism For CNS Penetration

**DOI:** 10.1101/2020.12.09.20242396

**Authors:** Carlotta Pipolo, Antonio Mario Bulfamante, Andrea Schillaci, Jacopo Banchetti, Luca Castellani, Alberto Maria Saibene, Giovanni Felisati, Maurizio Quadrio

**Affiliations:** Unit of Otolaryngology, ASST Santi Paolo e Carlo, Department of Health Sciences, Università degli Studi di Milano, Milan, Italy; Dept. of Aerospace Science and Technologies, Politecnico di Milano, Italy

**Keywords:** Computational Fluid Dynamics, SARS-Cov-2, Olfactory Mucosa, Nose

## Abstract

**Introduction:** SARS-CoV-2 is a respiratory virus supposed to enter the organism through aerosol or fomite transmission to the nose, eyes and oropharynx. It is responsible for various clinical symptoms, including hyposmia and other neurological ones. Current literature suggests the olfactory mucosa as a port of entry to the CNS, but how the virus reaches the olfactory groove is still unknown. Because the first neurological symptoms of invasion (hyposmia) do not correspond to first signs of infection, the hypothesis of direct contact through airborne droplets during primary infection and therefore during inspiration is not plausible. The aim of this study is to evaluate if a secondary spread to the olfactory groove in a retrograde manner during expiration could be more probable.

**Methods:** Four three-dimensional virtual models were obtained from actual CT scans and used to simulate expiratory droplets. The volume mesh consists of 25 million of cells, the simulated condition is a steady expiration, driving a flow rate of 270 ml/s, for a duration of 0.6 seconds. The droplet diameter is of 5 μm.

**Results:** The analysis of the simulations shows the virus to have a high probability to be deployed in the rhinopharynx, on the tail of medium and upper turbinates. The possibility for droplets to access the olfactory mucosa during the expiratory phase is lower than other nasal areas, but consistent.

**Discussion:** The data obtained from these simulations demonstrates the virus can be deployed in the olfactory groove during expiration. Even if the total amount in a single act is scarce, it must be considered it is repeated tens of thousands of times a day, and the source of contamination continuously acts on a timescale of several days. The present results also imply CNS penetration of SARS-CoV-2 through olfactory mucosa might be considered a complication and, consequently, prevention strategies should be considered in diseased patients.

## Introduction

SARS-CoV-2 is a respiratory virus, still widely spreading throughout the globe, that is presumed to enter the organism through aerosol or fomite transmission to the nose, eyes and oropharynx (1,2). Presentation ranges from respiratory symptoms including cough and fever to neurological symptoms like headache, dizziness and hyposmia, showing different target organs of the virus (3,4). Current literature has recently started to study access points into the CNS and the anatomical proximity between neurons, nerve fibres and the mucosa within the olfactory groove (5); the reported clinical-neurological signs related to alteration in smell suggest SARS-CoV-2 exploits this neuro-mucosal interface as a port of entry. Even though early reports (6) are indeed supporting this hypothesis through autopsy sampling, no literature exists as to how the SARS-CoV-2 reaches the mucosa at the level of the olfactory cleft, and whether the olfactory mucosa involvement is a direct consequence of viral particle deposition or due to a secondary viral invasion of these tissues during the infection.

From other respiratory viruses we know that aerosols, which are responsible for the transmission of airborne microorganisms, consist of small droplet nuclei (1–5μm) or droplets (>5μm) (7); these have specific characteristics regarding their distribution inside the nose and respiratory tract. Considering that hyposmia more commonly follow the first signs of presentation of infection (8,9), the hypothesis of direct contact through airborne at the stage of primary infection and therefore during inspiration is not plausible.

The second hypothesis of a secondary spread to the olfactory groove in a retrograde manner during for example expiration in an already challenged organism appears more likely (10). This would make CNS penetration a complication secondary to e.g. pulmonary infection, thus opening the field to so far unconsidered preventative measures.

Our group has therefore used computational fluid dynamics to study distribution of airflow and deposition of supposed infectious sub-micron droplets during breathing, to better understand the possible routes of infection and penetration inside the nasal cavity and the olfactory mucosa.

## Materials and Methods

This study was granted exemption from the Institutional Review Board of the San Paolo Hospital, Milano, Italy, due to its retrospective nature and is based on a set of CFD simulations of breathing, where only expiration is considered. The Large Eddy Simulation (LES) technique on CT scan reconstruction of nasal anatomy is used. LES is a high-cost and high-fidelity CFD approach, which allows fine control over the modelling error in dealing with complex and possibly turbulent flows. LES numerical simulations were performed starting from a set of four CT scans, whose sinonasal anatomy was defined by consensus by all authors as lacking any appreciable anatomic anomaly (i.e. a straight septum, normotrophic turbinates with orthodox bending, symmetrical distribution of anatomical features among the two sinonasal emi-systems).

CT scans have a 512 × 512 matrix with a 0.49 mm × 0.49 mm spatial resolution in the sagittal-coronal plane and a 0.625 mm gap between consecutive axial slices, with 250-350 native images for each case. More details on CT image processing, choice of the threshold value and 3D reconstruction, carried out via the software 3D Slicer (11), have been already reported in the literature documenting the entire procedure (12–16). The CFD simulations were carried out with the finite-volumes OpenFOAM software package (17).

The CFD analysis of each of the four cases (patients from P1 to P4) was conducted on a finely discretized volume mesh consisting of 25 million of cells, yielding extreme accuracy. The simulated condition is a steady expiration driving a flow rate of 270 ml/s, which corresponds to low to medium intensity breathing (18), for a duration of 0.6 seconds. A considerable number of droplets with a diameter of 5 μm, in accordance with the expected droplet size described above (9), were placed at the lower boundary of the computational domain and allowed to enter as time progresses. Droplets are transported by the airflow, and most of them are exhaled after travelling through the nasal chamber, becoming responsible for the potential airborne contagion. Due to their inertia, however, a fraction of the droplets deposits on the mucosal lining of the nose. The simulations identify the deposited droplets, and therefore provide a quantitative representation of the deposition pattern, highlighting areas of preferential deposition during expiration. Particular attention is given to the particles that reach the olfactory slit, qualitatively sketched in figure 1. This study provides information on the preferential site of adhesion of expiratory droplets to the olfactory mucosa and computes the spatially varying degree of probability for a droplet to deposit in a specific location instead of being convected to the external ambient.

**Figure 1.**
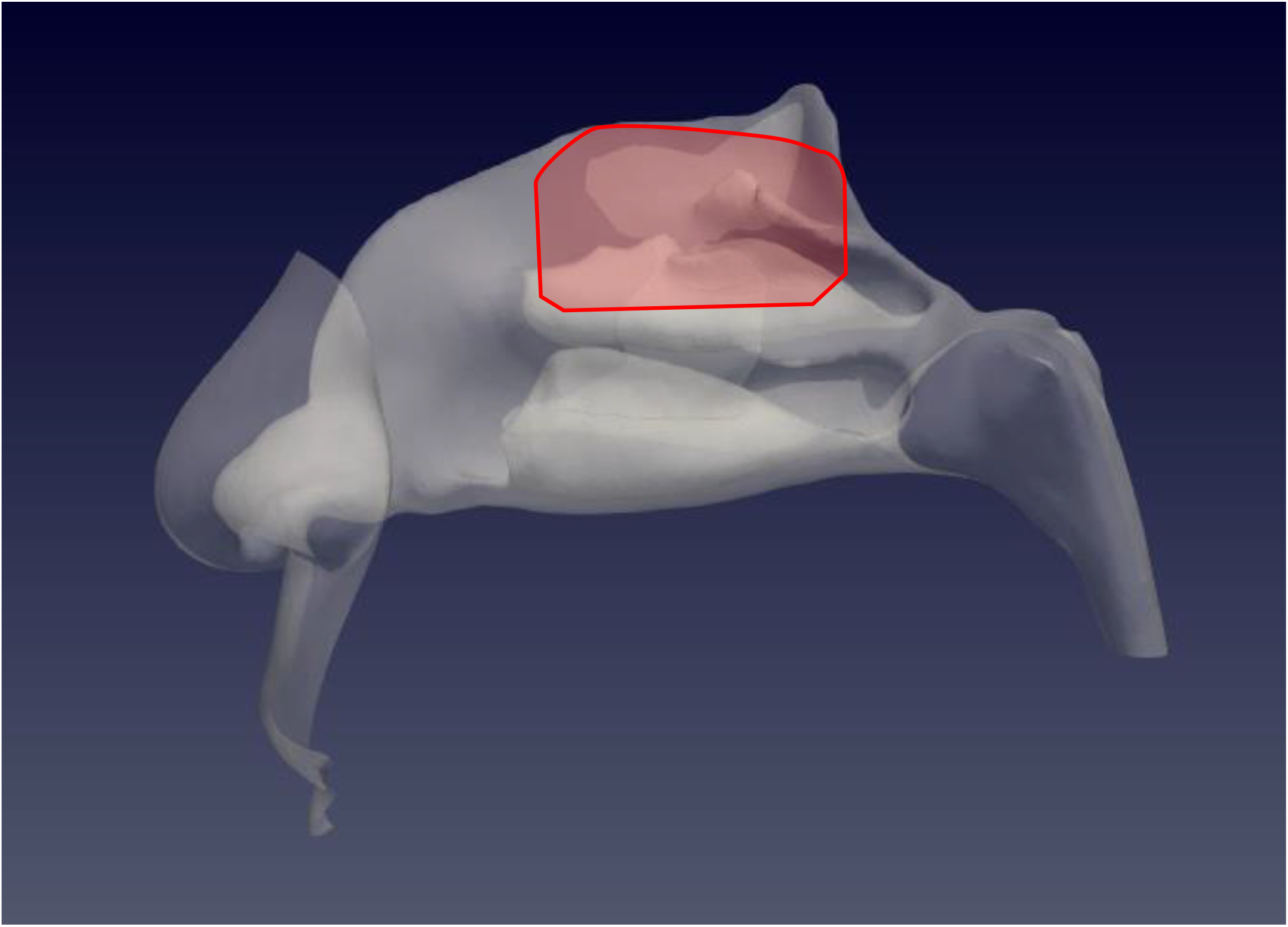
Sagittal view of the left nostril. The olfactory slit is highlighted in red.

## Results

The simulations portrait the preferential sites of droplet deposition on the nasal mucosa during expiration. It is clearly visible in Figure 2, 3 and 4 that, although virus deposition is prevalent in the nasal vestibule and rhinopharynx. Some droplets indeed do deposit in the area corresponding to the olfactory mucosa. Moreover, as expected, interindividual differences are visible. Droplets have been emphasized with a red dot.

**Figure 2.**
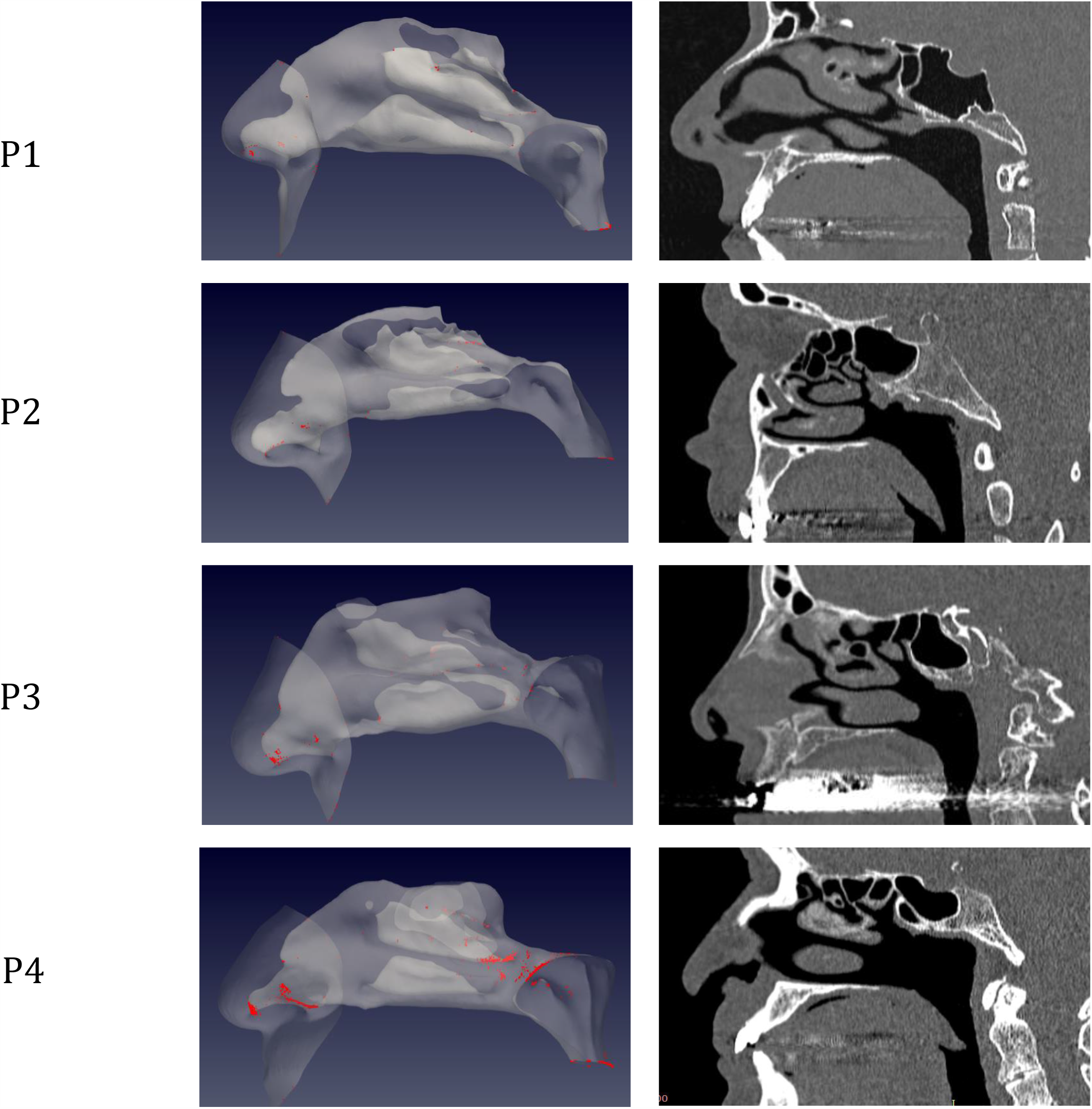
Distribution of droplet deposition during expiration, in sagittal projection. 3D models of the nasal fossae (left column), obtained from the CT, which are shown in the right panel. Particle size is increased to improve clarity.

**Figure 3:**
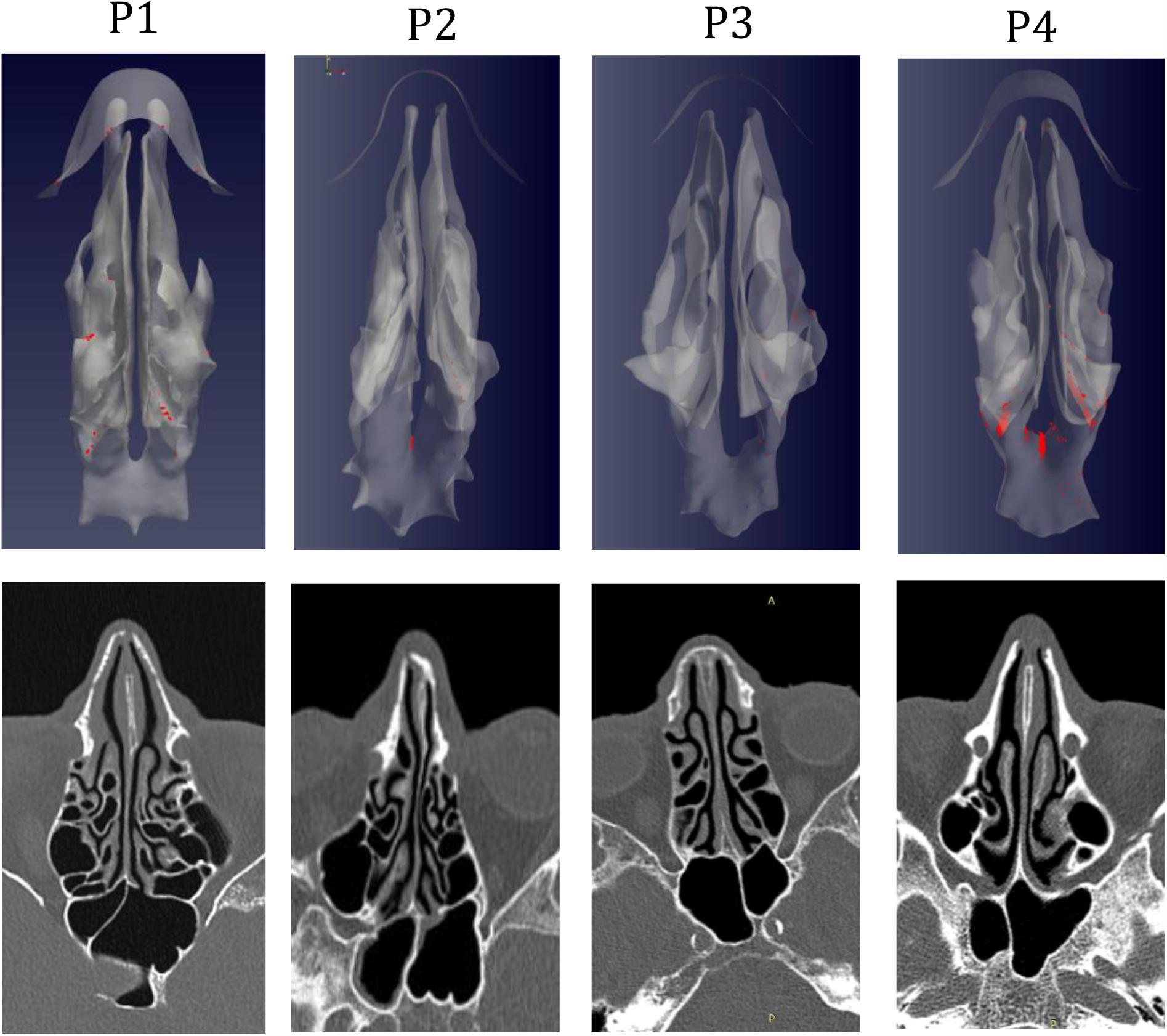
As in figure 2, but axial projection.

**Figure 4:**
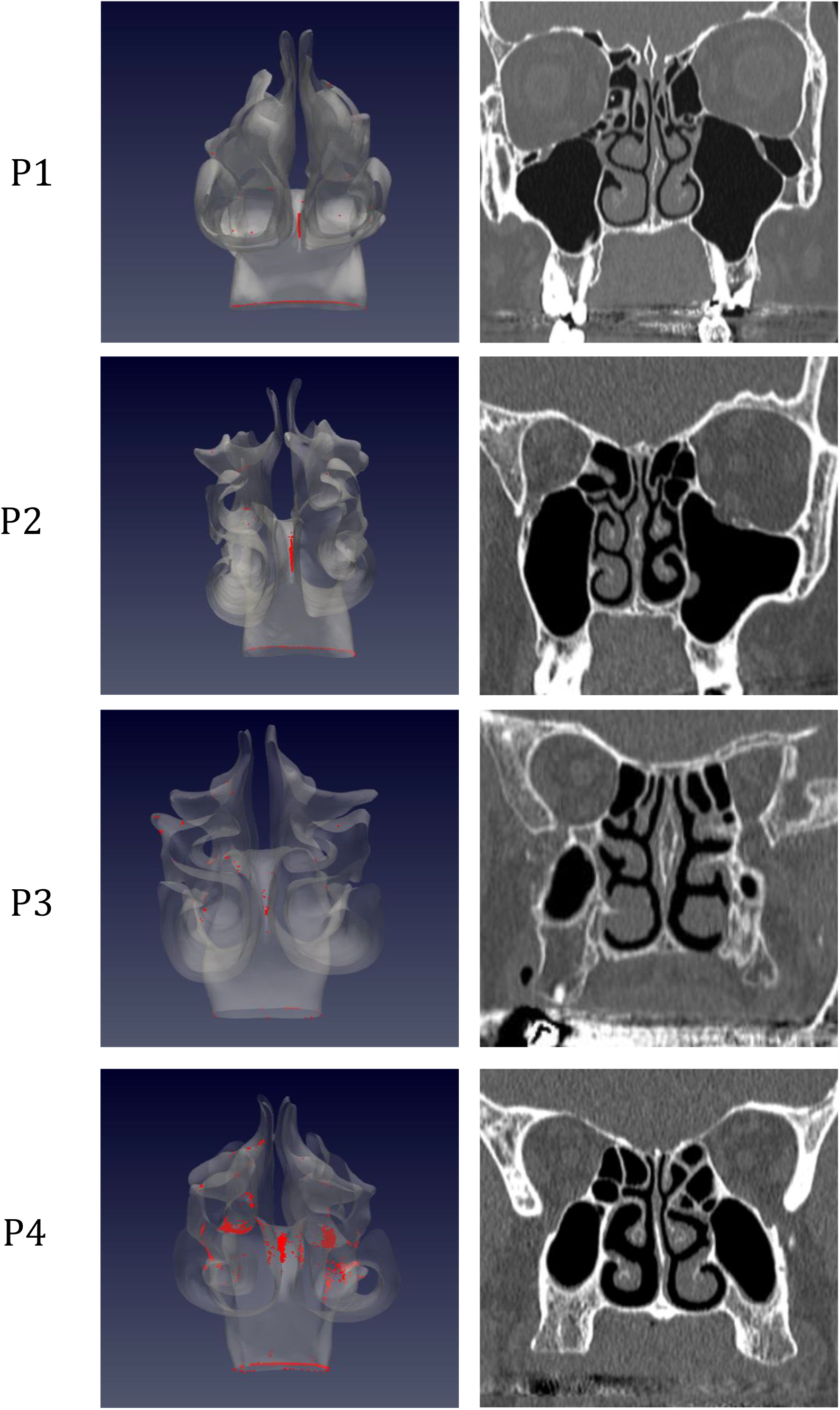
As in figure 2, but coronal projection.

The analysis of sagittal sections (fig. 2, where only the left nasal fossa is shown for clarity) shows the virus to have a high probability to be deployed in the rhinopharynx, on the tail of medium and upper turbinates. The possibility for droplets to access the olfactory mucosa during the expiratory phase is of primary interest. The evaluation of axial projections (Fig. 3) confirms a high concentration of particles in the posterior segments in addition to a better visualisation of particle distribution between medial and lateral compartments. Although heterogeneously, it can be observed how the particles, and consequently the virus, are more likely to settle in the medial quadrants of the nasal cavities than in lateral ones. During the expiratory phase, the particles have a significant probability to impact and adhere to the mucosa of the ethmoidal rostrum. Finally, the analysis of coronal projections (Fig. 4) confirms previous observations, although it demonstrates a better visualisation of the septal rostrum region and of both portions of the rhinopharynx. Coronal projections confirm indeed some particles reach the olfactory slit.

## Discussion

Regardless of the mechanism for viral transmission (direct respiratory, aerosol or fomite), first access to the nose must happen through inspiration. Once the virus has gained entry to the sinonasal cavity, however, many potential mechanisms concur to further diffusion, among which transport, local replication, and invasion of proximal structures. The ability of SARS-CoV-2 to bind the ACE-2 receptor, enter the respiratory epithelium cells and thereby initiate its replication has been thoroughly demonstrated (19,20).

Respiratory droplets containing viral particles are unable to massively reach the olfactory cleft, which should not be therefore considered a primary target for COVID-19 infection. The droplet ability to deposit on the olfactory cleft is a direct function of the particle size, given that the olfactory cleft is anatomically developed to receive smaller particles like odorants, while droplets carrying the viral load can be larger (10). Such an ineffective viral deposition onto the olfactory mucosa, coupled with the known defensive mechanisms employed by the olfactory mucosa to protect from environmental noxae (19,21), make the direct infection of the olfactory cleft by SARS-CoV-2 at the time of primary entry into the organism unlikely at best. Conversely, cumulative exposure of the olfactory cleft to expiratory droplets from the lower respiratory tract in an already diseased organism may be more likely. The viral load in the lung is much higher than a one-time aerosol reaching the nose, the act of expiration is repeated tens of thousands of times a day, and the source of contamination continuously acts on a timescale of several days. This route to the olfactory cleft and maybe to CNS may also explain the time lag between first symptoms and first neurological impairments including hyposmia (22,23).

The present results also imply CNS penetration of SARS-CoV-2 through olfactory mucosa might be a complication of an already present infection of the lower respiratory tract. Hence, prevention of the olfactory mucosa penetration by the virus should be considered in diseased patients. High volume nasal washes, usually performed with saline, can be used to reduce the adherence of viral parts emitted from the lower respiratory tract towards the nasal cavity, thus weakening the virus ability to spread to the olfactory mucosa. Indeed, other authors have advocated use of nasal lavages in SARS-CoV-2 infection as a preventive measure (24), and prior studies on viral upper respiratory tract infections with hypertonic saline showed reduced viral shedding and patient infectivity (25) in the already diseased. Prior studies on viral upper respiratory tract infections with hypertonic saline showed decreased viral shedding and reduced patients’ infectivity (25) in the already diseased. Others propose other types of medications: in (26) inhalation of acetic acid is suggested being effective in shortening the duration of symptoms. The present study -- besides suggesting the olfactory region as a target for inhibition of the secondary viral infection which endangers the CNS -- provides further support for the effectiveness of such preventive measures, since a diffuse droplet deposition takes place in the nasal fossae, that can be easily reached by washing or by other nasal medications.

Further studies are thus required to focus on nasal washes not only as a preventive measure to infection but also as a means to inhibit the secondary spreading of the virus to the olfactory mucosa and therefore to the CNS for COVID-19 patients. These studies should clinically quantify the ability of nasal washes, i.e. a simple and non-invasive treatment, to halt the progression of the disease by containing its complications involving the CNS.

## Data Availability

Results are available on request

## Abbreviations

SARS-CoV-2: severe acute respiratory syndrome coronavirus 2
CT: computed tomography
CNS: Central nervous system
LES: Large eddy simulation
CFD: Computational fluid dynamics
ACE-2: Angiotensin converting enzyme
COVID-19: Coronavirus disease 2019

